# Treatment interventions and assessment of fear-avoidance for chronic musculoskeletal pain: a systematic review protocol

**DOI:** 10.1101/2024.06.03.24308385

**Authors:** S. Tan, A. Jaggi, A. Tasker, C. Borra, F. Watson

## Abstract

**Introduction:** The Fear-Avoidance (FA) model aims to explain how an acute pain experience can develop into a persistent state. The FA model considers five core components: kinesiophobia, pain-related fear, catastrophisation, victimisation, and interpersonal social environment. Amongst these, kinesiophobia, tends to dominate the literature on chronic musculoskeletal pain. As a result, current reviews have not considered the other core components of the FA model when exploring its interventions. Moreover, several synonyms of the term kinesiophobia is not reflected in their search strategies. Coupled with the preference of particular study designs and outcome measures, this systematic review aims to provide and characterise an overview of treatment interventions that consider all study designs, relevant outcome measures, FA components, and FA component synonyms.

**Methods and analysis:** Eligible studies will be in English or with an available English translation from 1970 onwards. Databases to be searched include Cochrane Central Register of Controlled Trials (CENTRAL), MEDLINE, Embase, The Allied and Complementary Database (AMED), PEDro, Web of Science, and grey literature. We will include studies involving participants ≥18 years old with chronic musculoskeletal pain, and interventions targeting FA and/or its components. Three review authors will independently screen papers using preestablished eligibility criteria and conduct assessments of risk of bias, with a fourth independent researcher employed to resolve disagreements where found. Qualitative synthesis techniques will be used to characterise the interventions. Patient and Public Involvement (PPI) has been utilised to develop this protocol and will be conducted following completion of the systematic review to discuss and reflect on the findings.

**Ethics and dissemination:** This systematic review does not require ethical approval as existing data will be used and the PPI to be conducted is an involvement activity rather than study data. The results will be disseminated through a peer-reviewed journal and via national and international conferences.

**Prospero registration number:** [this protocol will be registered following peer review].

**Strengths and limitations of this study:**

- This systematic review provides a meaningful and patient-centred search in the context of fear-avoidance (FA), as the search strategy purposively includes additional components of FA and its synonyms.
- The search strategy will include eight relevant measures and all study designs to provide a diverse overview of the current interventions for FA in chronic musculoskeletal conditions.
- Utilising a combination of established qualitative methods to identify and characterise interventions for fear avoidance and integrating them with the opinions of the PPI community collaborators to better appreciate multiple perspectives and inform further research.
- A meta-analysis is not considered due to the methodological heterogeneity of including all study designs, however it is not entirely applicable to the scoping objective of this systematic review.
- The inclusion of participants with comorbidities increases generalisability of results at the risk of introducing additional biases and confounders.

## Introduction

### Rationale

The Fear-Avoidance (FA) model was developed by Lethem et al in 1983 (26) to explain the pathways by which acute pain transitions to chronic using a biopsychosocial perspective (1, 2). According to the FA model, a self-reinforcing cycle of catastrophising, fear, hypervigilance, and further avoidance of movement can lead towards dysfunction, deconditioning, depression, and increased pain (3). The FA model is made up of four main components: kinesiophobia, catastrophising, pain-related fear, and FA beliefs (4, 5). Some researchers have suggested adding victimisation, disability, self-efficacy, and input from the interpersonal social environment, suggesting that the model should extend beyond pain-related fear and highlights the significance of concurrent, and often competing, goals (2, 6-8) (Fig 1).

**Fig 1:**
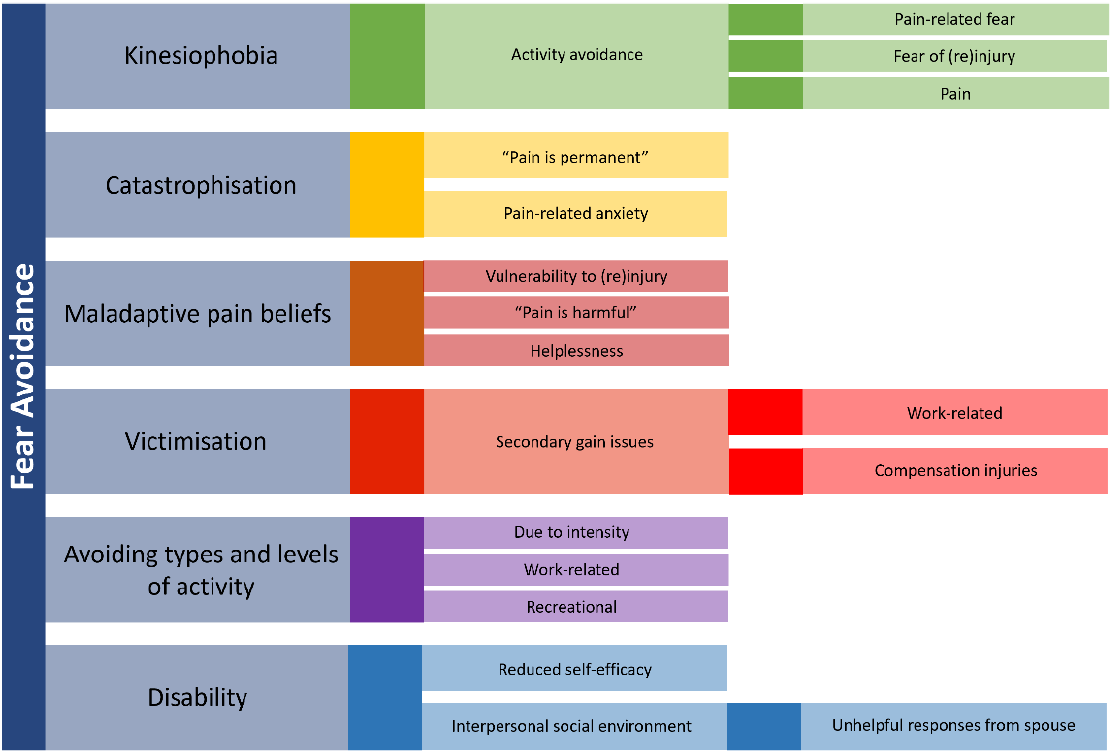
The components of the Fear Avoidance model and potential implications.

In the assessment and treatment of chronic musculoskeletal pain, kinesiophobia may be described and understood by a range of synonyms (e.g., “fear of movement”, “fear of (re)injury”, “avoidance behaviour”, “avoidance hypervigilance”, “behavioural performance”, “pain-related fear with impaired physical performance”, and “avoidance of activity”) which tend to dominate the research landscape (9-11). Kinesiophobia is an important mediator of new and prolonged chronic pain and disability, and as such is an important element of the FA paradigm; rates are estimated at 51-72% of people with chronic pain (9, 12). Kinesiophobia presents a major challenge to successful physical rehabilitation; unchecked, it may contribute to the development of disuse syndrome, which is associated with additional physical and psychosocial impacts such as depression, muscle atrophy, and medication misuse (13). These fundamental effects risk drawing practitioners focus away from less-reported aspects of the FA model.

Several systematic reviews have focused on the results of interventions to treat kinesiophobia (14-17); a focus which risks overlooking the multifactorial origins and drivers of chronic musculoskeletal pain. Bordeleau et al (9) recommends further research into the multi-faceted nature of kinesiophobia, encompassing physical, psychological, and social interventions. These concerns are further compounded by the nature and use of tools available to measure kinesiophobia; methods such as the Tampa Scale of Kinesiophobia (TSK) that is widespread and accessible (18). Liu et al conducted a review of related measures and found that The Fear-Avoidance Beliefs Questionnaire (FABQ) and TSK were the most often used questionnaires (19,20). Indeed, many systematic reviews have specified use of the TSK as part of their study inclusion criteria (9, 15). This further exacerbates the focus on kinesiophobia, and simultaneously dampens potential for assessment of the full FA model on chronic musculoskeletal pain. Moreover, current reviews have primarily included randomised controlled trials (RCT) in their search strategy, which restricts the diversity of interventions explored.

In addition to the TSK and FABQ, there are other FA model-adjacent measures including the Kinesiophobia Causes Scale (KCS) (21), Athlete Fear-avoidance Questionnaire (AFAQ) (5), and the Fear-Avoidance Components Scale (FACS) (7). Some researchers have started to use a combination of the aforementioned measures, but it is unknown whether this is appropriate given the potential for discordance between each measure and the FA model construct (9, 22-24).

The current literature on assessing FA and its components, beyond just kinesiophobia, is lacking. There is a need for a systematic review that includes intervention studies assessing and/or treating FA by appreciating its components on chronic musculoskeletal pain, as it is more patient-tailored and clinically useful. As such, utilising this study’s interpretation of the FA model will influence the search criteria to include all relevant study designs and the current measures of FA components. Finally, the voice of community collaborators, such as patients, carers, and allied health professionals is vital in ensuring patient-centred conclusions and recommendations for assessment and interventions of FA in musculoskeletal health. Therefore, the objectives of this systematic review are as follows:

1. Identify and characterise interventions used to treat FA in people with chronic musculoskeletal pain by considering all study designs, relevant measures, and FA synonyms;
2. evaluate the use of common FA model-related measures that assess treatment interventions in people with chronic musculoskeletal pain;
3. engage with relevant community collaborators in the development of the systematic review search methodology and, at completion, to provide reflections on the findings and ensure further research to be clinically relevant.

## Methods

This protocol will follow the Preferred Reporting Items for Systematic Reviews and Meta-Analysis Protocols (PRISMA-P) reporting guidelines (25). This study is registered with PROSPERO (INSERT ID FOLLOWING PEER REVIEW) and will be conducted between 1^st^ July 2024 – 1^st^ September 2024.

### Patient and Public Involvement Statement (PPI)

A panel of patients with chronic pain who attended the Royal National Orthopaedic Hospital, UK attended an online focus group about chronic pain and were reimbursed for their time. The nature of this PPI is an involvement activity, where the opinions of community collaborators were collected on the topic of FA, and so ethical approval is not required. Regarding kinesiophobia, patients spoke about the importance of early intervention, education, and rehabilitation from encouraging, knowledgeable, and positive health professionals. Within the discussion, participants mentioned other aspects of fear avoidance, such as catastrophisation, and were concerned that their fear of movement would never fully disappear but felt hopeful that it could be dampened with correct management. Participants all agreed that peer support was vital to their personal ‘journey’ in managing fear of movement. Moreover, a concern was raised about how a participant felt that they were unable to take part in research studies due to their comorbidities, and so consequently believed that their views and experiences were not reflected in the research.

The authors felt that the term ‘kinesiophobia’ was quite uncommon among allied health professionals, musculoskeletal researchers, and patients, and that research may exist on this topic without naming it in such a way, and consequently be missed by systematic review searches. To combat this, the aim was to reach multiple health professionals to consider their understanding of kinesiophobia so that they could offer the terminology they use with other clinicians or their patients. The authors circulated the following question on X^i^ (formerly Twitter),

> *Calling ALL health professionals! What do you mean when you say *kinesiophobia*? Please comment below + include a description of your job role + share with your network*.

This resulted in 20,001 impressions, 727 engagements, 23 retweets and 37 comments. Direct responses came from 36 individuals, representing physiotherapists, musculoskeletal researchers, orthopaedic surgeons, pain management clinicians, chiropractors, and sports trainers. Examples of responses include,

> *“…being scared to move too much for fear of triggering a flare up of pain*.*” “…rationale of irrational fear of causing or increasing tissue damage…”*
>
> *“when an individual doesn’t have the knowledge, skills and/or confidence to move their body…”*
>
> *“I don’t tend to use the word clinically however, in children+young[sic] people I would describe kinesiophobia as the loss of spontaneous, fluent and playful movement due to a memory or thought that links movement to a negative and feared consequence*…*”*
>
> *“…no patient has ever used the term unless learnt from a clinician*.*”*
>
> *“…I don’t like when it’s describe[sic] as “irrational” fear it makes me feel like we’re blaming the person*.*”*
>
> *“…delineating terms may help us work more effectively with patients…”*

These interactions subsequently informed the design of our search terms and inclusion/exclusion criteria.

In addition to this protocol being informed by PPI, results of this systematic review will be shared with another focus group including patients, carers, and health professionals to discuss their impression of the findings and potential for impact on them, to gauge how well their experience is represented by the findings and generate ideas for future research.

### Eligibility criteria

Eligible studies include primary interventional research articles in English or with an available English translation. The FA model was initially outlined by Lethem et al (26) and studies that explored psychological factors in chronic pain were increasingly prevalent in the 1970s, so the date range for included reports will be from 1970 onwards.

### Information sources

Cochrane Central Register of Controlled Trials (CENTRAL), MEDLINE, Embase, The Allied and Complementary Medicine Database (AMED), PEDro, Web of Science, and grey literature (e.g., medRxiv, ISRCTN protocol registry) will be searched using database-specific syntax detailed below. Reference lists of resulting articles will be searched for articles previously missed. The final list of selected articles will be entered into Connected Papers^ii^, which will also search for associated papers that could have been missed.

### Search strategy

The search strategy will be formulated using the Peer Review of Electronic Search Strategies (PRESS) Evidence-Based Checklist (27). The searches will be divided into three themes: [1] musculoskeletal conditions, [2] FA components and their synonyms, and [3] relevant outcome measures. These themes will be combined with the Boolean operator “AND”. Any restrictions outlined in the exclusion criteria will be combined cautiously, as to not remove relevant articles, with “NOT”.

Ovid^iii^ will be used for the AMED, Embase, and MEDLINE databases whilst CENTRAL and PEDro will be searched on their own respective search engines. Key search terms will be used for every database and logically matched with their respective subjecting headings or Medical Subject Headings (MeSH) term system. For instance, Ovid has a “Map Term to Subject Heading” option, whilst CENTRAL uses MeSH terms. The relevant subject headings are database-specific and will be exploded where necessary. If a subject heading system is unavailable, then a multi-purpose (mp) search would be used for the free-text key terms, i.e. several fields including title, abstract, and keywords (27).

The key search terms for the search strategy are:

1. **Search 1 (musculoskeletal conditions):** “pain”, any MSK “-algia” conditions (e.g. causalgia, neuralgia, epicondyalgia, etc.), MSK pain, physical suffering, MSK pain (any body part, e.g. elbow, knee, neck, back, etc.), “phantom limb”, “complex regional pain syndrome”, “sciatica”, “neuropath*”, arthritis (all forms, e.g. osteoarthritis, rheumatoid arthritis, etc.), “sacroilitis”, “Failed back surgery syndrome”, “persistent spinal pain syndrome”, “whiplash”
2. **Search 2 (FA components):** kinesiophobi*, “fear of mov*”, “fear of injur*”, “fear of reinjur*”, “fear of re-injur*”, “fear” AND “movement”, “fear” AND “injur*”, “fear” AND “reinjur*”, “fear” AND “re-injur*”, “fear avoidance”, “behavioural performance”, “behavioral performance”, “avoidance behaviour”, “avoidance behavior”, “pain-related fear”, “avoidance of physical exertion”, “avoidance hypervigilance”, “impaired physical performance”, “scared of movement”, “mov* anxiety”, “avoidance learning”, “catatastrophi?*”, “maladaptive pain belief”, “pain belief”, “victimi?ation”
3. **Search 3 (relevant outcome measures):** “ Tampa Scale of Kinesiophobia”, “Kinesiophobia Causes Scale”, “Fear-Avoidance Beliefs Questionnaire”, “Fear-Avoidance Components Scale”, “Athlete Fear-Avoidance Questionnaire”, “Pain Anxiety Symptoms Scale”, “Pain Catastrophi?ing Scale”, “Patient-Reported Outcomes Measurement Information System Pain Interference”
4. **Search 4:** Search 1 AND Search 2 AND Search 3
5. **Search 5: Search 4 NOT exclusion criteria (e.g. postoperative populations)**

Table 1 shows an example search of the MEDLINE database using the Ovid platform:

**Table 1.**
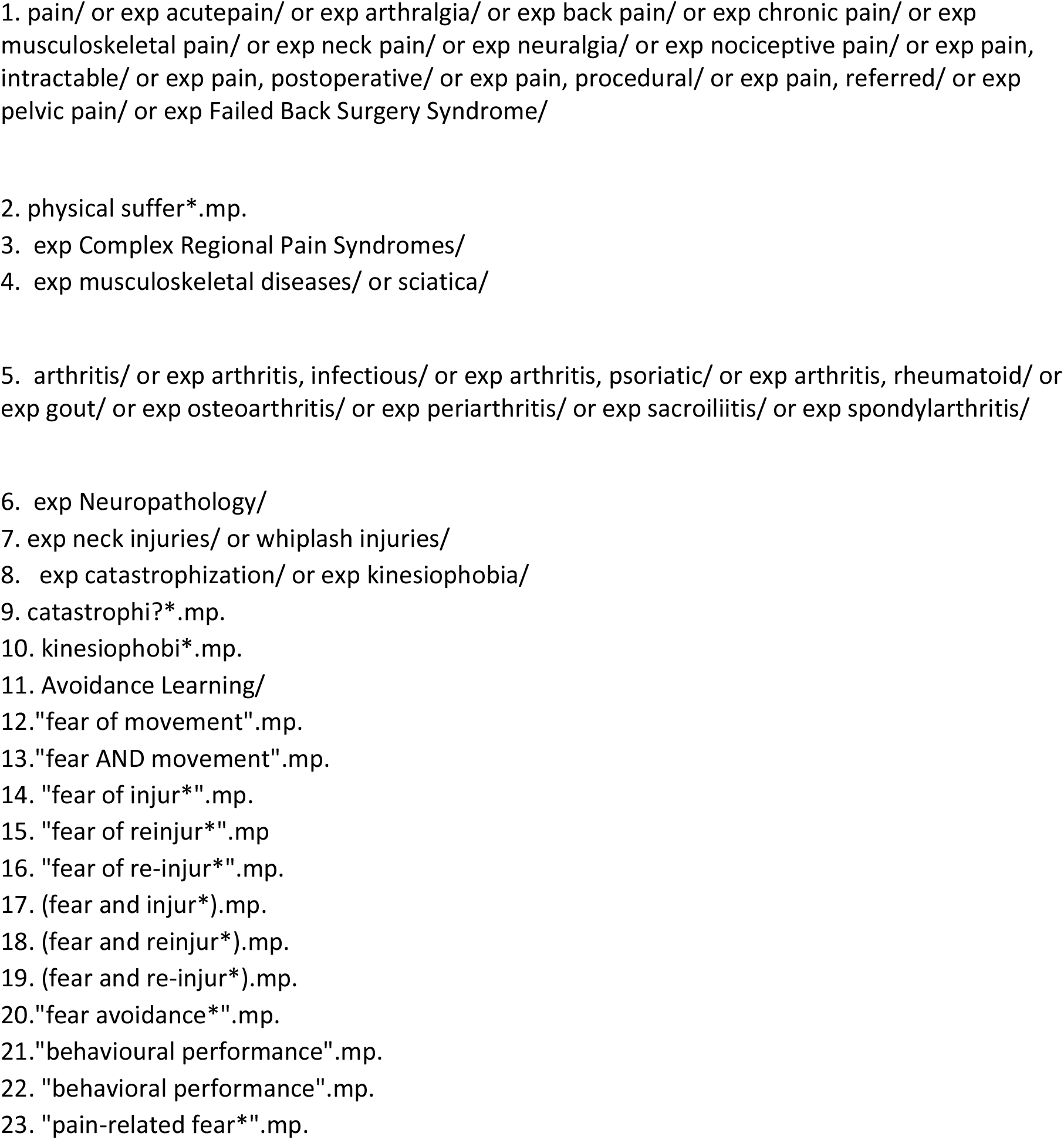
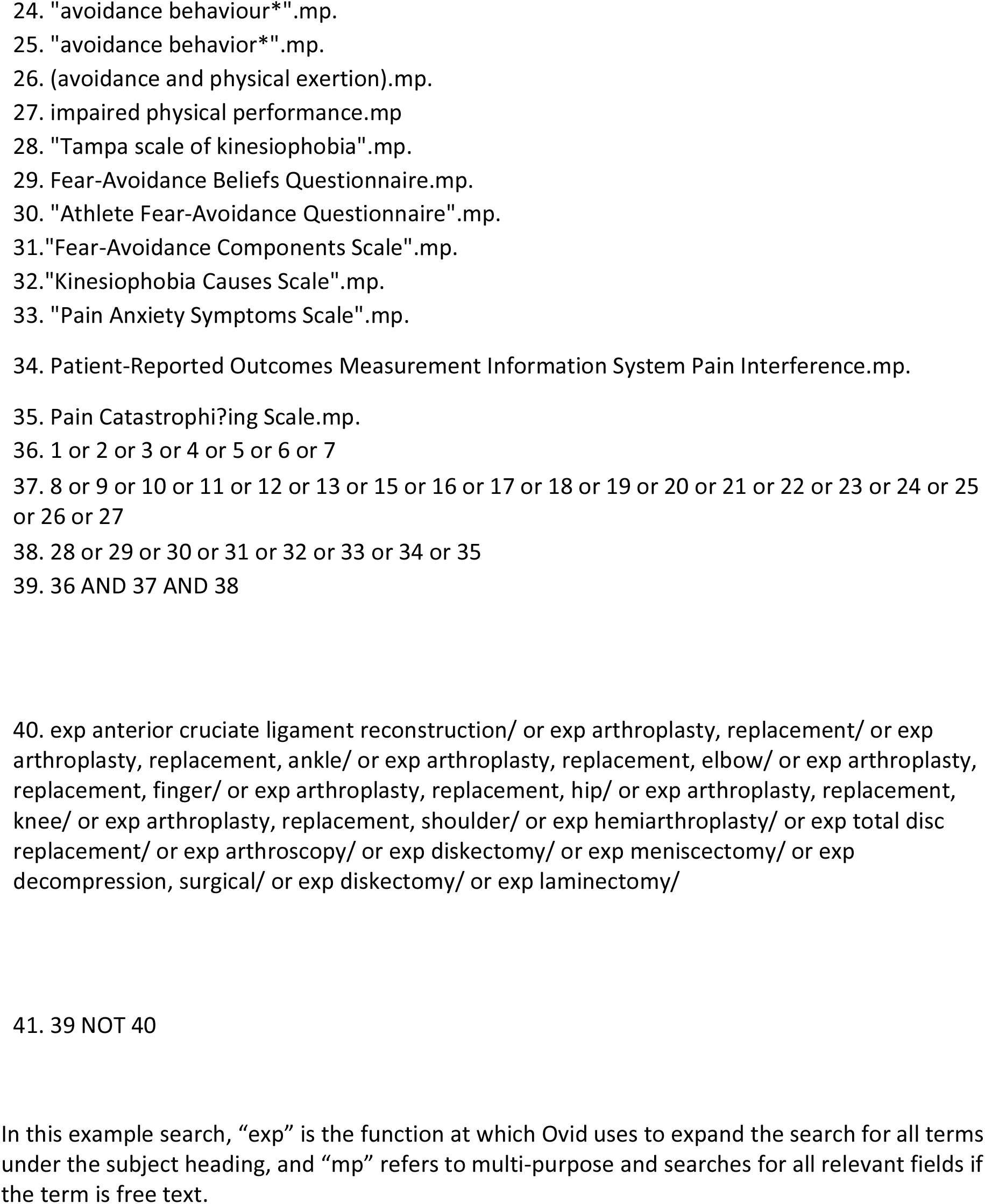
Example search of the MEDLINE database on Ovid.

### Study records

#### Data management

Search results will be exported from the database and compiled in Rayyan^iv^. Rayyan is an online tool that is designed for screening reports in systematic reviews and allows convenient collaboration amongst the researchers, such as with remote working. There is also an additional feature that allows the automatic detection and removal of duplicates, which will be done at the 95% confidence threshold within the Rayyan settings.

#### Selection process

Using Rayyan, three researchers including clinicians and academics (ST, FW, AJ) will independently review the search results to deem whether studies meet predetermined inclusion/exclusion criteria using the title and abstract. The full text of remaining studies will be reviewed independently to determine final inclusion in the systematic review. Where disagreements occur, an impartial researcher (AT) will help reach resolution through group discussion. A flow diagram will be produced showing papers excluded at each stage, along with the reason.

#### Inclusion criteria

Papers describing individuals of any sex who are aged ≥18-years old and targeting chronic pain resulting from any musculoskeletal condition will be included in this study. Included studies can contain homogenous or heterogenous populations and individual participants can experience multiple comorbid conditions. Any intervention (e.g. physical, psychiatric, educational, psychosocial, behavioural) will be included with or without comparison to a control group, and regardless of outcome. The intervention must target FA and/or its components by the context of the study. The questionnaire used to assess FA and/or its components can either be primary or secondary outcome. The included questionnaires used to measure fear avoidance are outlined in search 3 of the above search strategy. Studies will be considered regardless of setting, time frame, or treatment length. No limits nor filters will be used to restrict the study designs included.

#### Exclusion criteria

Studies on children, solely healthy subjects, non-musculoskeletal fear avoidance (e.g., neurological origin in people with Parkinson’s or Multiple Sclerosis, or visceral pain), and postoperative populations (e.g. following total knee arthroplasty) will be excluded. Review articles and secondary analysis papers will be excluded.

#### Data collection process

A data collection form detailing all data items will be used to extract information for included papers by a single researcher. Where data is missing, two attempts will be made to contact the corresponding author of the papers at two-week intervals, to retrieve the necessary information.

### Data items

Study identifiers, study design, cohort size, cohort location, intervention, participant gender, participant age, cohort diagnosis/disorder, cohort symptom characteristics, cohort comorbid diagnoses, treatment period (e.g., 6-months), outcome measure (e.g., TSK), treatment outcome, comparative group (where applicable), power (where applicable and achievable), effect size (where applicable and achievable), level of evidence for therapeutic studies (28).

### Outcomes and prioritisation

The primary outcome will be a list of the identified and characterised treatment interventions used to treat FA components and the questionnaires used to assess outcome. Secondary outcomes will be the opinion of community collaborators through PPI to provide reflections and develop a list of future research priorities.

### Risk of bias of individual studies

For RCTs, the Risk of Bias (ROB) 2.0 tool (29) will be used to assess the risk of bias. For non-randomised studies, the Risk of Bias In Non-randomised Studies – of Interventions (ROBINS-I) tool (30) will be used. This tool considers that each study is attempting to mimic an RCT and judges them on similarly thorough criteria in sevens domains that could affect bias. Analysis of bias will be carried out for all included studies twice between two reviewers (ST, FW) independently. Where disagreements occur, discussion led by a mediator (AT) will aim to resolve differences and result in agreement. Both tools utilise relevant cut-off points that categorises the risk of bias as either “low risk of bias”, “some concerns”, or “high risk of bias”.

### Data synthesis

The identification and extraction of core themes from included papers will be performed using a combined reflexive thematic analysis and realist synthesis approach (32-35). Reflexive thematic analyses and associated thematic networks are established qualitative analytical tools designed to reveal and organise themes in a hierarchical manner through a structured approach. The themes generated during reflexive thematic analysis will be tested and interpreted by key community collaborators in the PPI groups; these interpretations will be integrated with the reflexive thematic analysis data through and adapted realist synthesis methodology. Realist synthesis has been applied to systematic reviews to uncover the mechanisms underpinning interventions positioned within contextual factors. This combination of established qualitative approaches will enable the robust integration of the systematic review and PPI reflexive data to better appreciate multiple perspectives on interventions.

### Meta-biases

Tabulation and visual display of the individual risk of bias assessments will be created. Due to the assumed heterogenous nature of the gathered studies, it will not be feasible to conduct a meta-analysis.

### Confidence in cumulative evidence

The Grading of Recommendations Assessment, Development and Evaluation (GRADE) method will be used to assess the quality of evidence for all outcomes (35). This includes risk of bias, consistency, directness, precision, and publication bias. This will result in allocation of a high, moderate, low, or very low-quality study label.

## Discussion

This systematic review will provide and characterise a broad overview of interventions intended to treat FA and evaluate the questionnaires used to assess their outcomes. Until now, many systematic reviews have focused on kinesiophobia as a single aspect of the FA model, rather than a more holistic approach, as is proposed here. A broad perspective of treatment interventions for FA related to chronic pain in musculoskeletal conditions could educate medical professionals and highlight areas of further research. Understanding how the outcome of these studies are assessed using questionnaires will allow recommendations to be made for future research and a stronger inclusion of the whole FA model rather than purely kinesiophobia.

A limitation of this protocol is the inclusion of participants with comorbidities because FA often results from, or is commonly comorbid with, other diagnoses. Whilst this will increase the risk of bias, it will ensure results are more generalisable (36), in keeping with our PPI focus group findings. Additionally, all types of study are included, rather than just RCTs, so assessment of risk of bias will be an important factor when drawing conclusions from this study, but we felt it was important to give a broad perspective at this stage.

In carrying out this work, we also hope to introduce an interesting PPI framework that could be used and expanded upon in future systematic review protocols. Public involvement could empower people with chronic musculoskeletal pain to take ownership of their treatment and seek out knowledgeable professionals who deliver evidence-based care. Additionally, it will ensure our research aims, perspectives and priorities are informed and directed by the exact population we aim to help.

## Data Availability

No datasets were generated or analysed during the current study. All relevant data from this study will be made available upon study completion.

## Footnotes

i www.twitter.com

ii https://www.connectedpapers.com/

iii https://ovidsp.dc1.ovid.com/ovid-new-a/ovidweb.cgi

iv https://www.rayyan.ai/

